# Raman spectroscopy on blood serum samples of patients with end-stage liver disease

**DOI:** 10.1101/2021.06.06.21258433

**Authors:** Staritzbichler René, Hunold Pascal, Estrela-Lopis Irina, Peter W. Hildebrand, Isermann Berend, Kaiser Thorsten

## Abstract

Raman spectroscopy has shown to be a promising method for the examination of biomedical samples. However, until now, its efficacy has not been established in clinical diagnostics. In this study, Raman spectroscopy’s potential application in medical laboratories is evaluated for a large variety (38) of biomarkers. Given 234 serum samples from a cohort of patients with different stages of liver disease, we performed Raman spectroscopy at 780nm excitation wavelength. The Raman spectra were analyzed in combination with the results of routine diagnostics using specifically developed complex mathematical algorithms, including fluorescence filtering, frequency subset selection and several overfitting circumventing strategies, such as independent validation. With the results of this cohort, which were validated in 328 independent samples, a significant proof-of-concept study was completed. This study highlights the need to prevent overfitting and to use independent data for validation. The results reveal that Raman spectroscopy has high potential for use in medical laboratory diagnostics to simultaneously quantify multiple biomarkers.

## Introduction

To diagnose, choose adequate therapy, and monitor the course of severe diseases, most patients require laboratory diagnostics. Individual laboratory tests are ordered specifically for the medical indication (targeted diagnostics). However, many different biomarkers are available, and selecting the appropriate diagnostic parameter can be challenging. A non-targeted diagnostic approach that could provide information on various relevant clinical conditions simultaneously could reduce the risk of missed diagnostics and delayed medical consequences.

Raman spectroscopy (RS) is a method capable of analyzing complex molecular compositions. It was first predicted by Adolf Schmelka in 1923 [1] and was first observed in 1928 by two groups of scientists in parallel [2], [3]. C.V. Raman received the Nobel price in 1930 for showing that a fraction of monochromatic light changes its frequency when traversing a transparent material [4].

RS is based on the inelastic scattering of photons due to their interactions with vibrational modes of molecules. This represents a principle distinct from absorption spectroscopy, which is based on transitions between the allowed energy levels of molecules. RS is related to infrared spectroscopy (IR) in that both reflect the internal oscillations within molecules. However, IR intensity reflects changes in dipole momenta, while RS reflects changes in polarization due to oscillations. IR and RS return different distributions of intensities and therefore provide complementary information.

RS is a powerful non-targeted analysis tool with numerous applications within and outside the clinical context. Airport security controls, for example, use RS, as it reveals even small traces of hazardous substances [5]. Another common application is in evaluating paintings’ originality [6]. Several diagnostic applications have been reported: [7], [8], also in the context of cancer: [9], [10], [11], [12], and in obesity-related diseases: [13], [14], [15]. Other studies deal with specific applications using blood: [16], [17], [18], [19], or molecular imaging: [20], [21]. Theoretically, due to the unique signatures of substances, countless biomarkers and metabolites could be analyzed in parallel within about one minute, enabling a fast medical response. Currently, laboratory diagnostics is based on individual chemical detection reactions, each requiring sample volume, reagents, and complex infrastructure. In contrast, a multi-target, reagent-free method could provide a complex clinical picture of the patient in a single step.

The aim of our study is to evaluate the applicability of RS as diagnostic tool for clinical samples in comparison to the results of routine medical laboratory diagnostics.

Due to its highly complex composition, blood analysis by means of RS is rather ambitious. Whole blood consists of cells, coagulation factors, and serum. The cells include red and white cells and platelets. The intracellular composition differs fundamentally from that of the cell-free blood components. Consequently, direct analysis of whole blood with RS may be extremely challenging. To reduce the analytical complexity, we used blood serum, which contains only cell-free components of the blood after the consumption of coagulation factors.

We used material available from a recently published study of patients with liver disease [22]. The cohort consists of patients at different stages of liver disease, from very early to the most advanced stages. Due to the dramatic disturbance of the metabolism, especially in patients with end-stage liver disease, this cohort provides a promising starting point for determining correlations between RS spectra and laboratory results. The clinical parameters of the patients are displayed in Table 1. The glomerular filtration rate (GFR) is a measure of renal function that includes creatinine, gender, and age. It was estimated using the CKD-EPI creatinine equation [23].

**Table 1.** Patient characteristics. (LC: liver cirrhosis.)

|  |  |
| --- | --- |
| Samples [n] | 234 |
| Number of patients | 137 |
| Sex [n] | 86male, 51 female |
| Age mean [years](range) | 56 (27 - 73) |
| Etiology of liver disease [n] |  |
| Ethyltoxic LC | 151 |
| Kryptogen | 22 |
| Hepatitis C | 16 |
| Hepatitis B | 5 |
| NASH | 12 |
| AIH | 6 |
| PSC | 4 |
| PBC | 6 |
| Laboratory diagnostics | mean (range) |
| MELD Score | 16 (6 - 39) |
| MELDNa Score | 15 (6 - 40) |
| Bili [$\mu\text{mol/l}$] | 48,8 (3.5 - 454.2) |
| INR | 1,5 (0.9 - 4.7) |
| Crea [$\mu\text{mol/l}$] | 91 (40 - 644) |
| GFR [$\text{mL/min/1.73 m}^2$] | 71,9 (7.8 - 134.7) |
| Sodium [$\text{mmol/l}$] | 137 (120 - 148) |

In our cohort, we focused especially on laboratory parameters for liver disease. Consequently, we included bilirubin, which is a routinely used detoxification parameter, and albumin, prealbumin, and cholinesterase as liver synthesis parameters. Furthermore, we included transaminases (ALAT, ASAT), alkaline phosphatase (AP), and *γ*-glutamyltransferase (GGT) as diagnostic biomarkers for injury of hepatocytes and alpha-fetoprotein (AFP) as a tumor marker for hepatocellular carcinoma. Model of end-stage liver disease scores (MELD, MELDNa) were analyzed, as these provide further information about patients’ prognosis and stage of liver disease. The MELD scores were calculated from the serum creatinine, serum bilirubin and prothrombin time values according to the applicable specifications in Europe [24]. The MELD-Na score incorporates the sodium blood concentration in addition to the MELD parameters and is used in the United States of America [25]. Other diagnostic laboratory parameters were also analyzed. We included creatinine and cystatin C as markers of renal function and C-reactive protein (CRP) and interleukin 6 as biomarkers of inflammation. Some general parameters, such as electrolytes (sodium, chloride, and phosphate), total protein, total cholesterol, ferritin, and iron were also included.

We also included the patients’ results for the following biomarkers in our analysis, although these were measured as whole blood samples or hemolysate and are not detectable in serum: glycated hemoglobin (hemoglobin A1c, HbA1c), blood cells (platelets, white blood cells), and international normalized ratio (INR)). Although serum samples should provide a promising starting point for determining the correlations between RS spectra and laboratory results, they posed some challenges. Specifically, the fact that biomarkers are naturally in solution poses a major inherent technical challenge – most characteristics found in the spectra of dry counterparts of these substances simply flatten out (see Figure 1 and 2).

**Fig 1.**
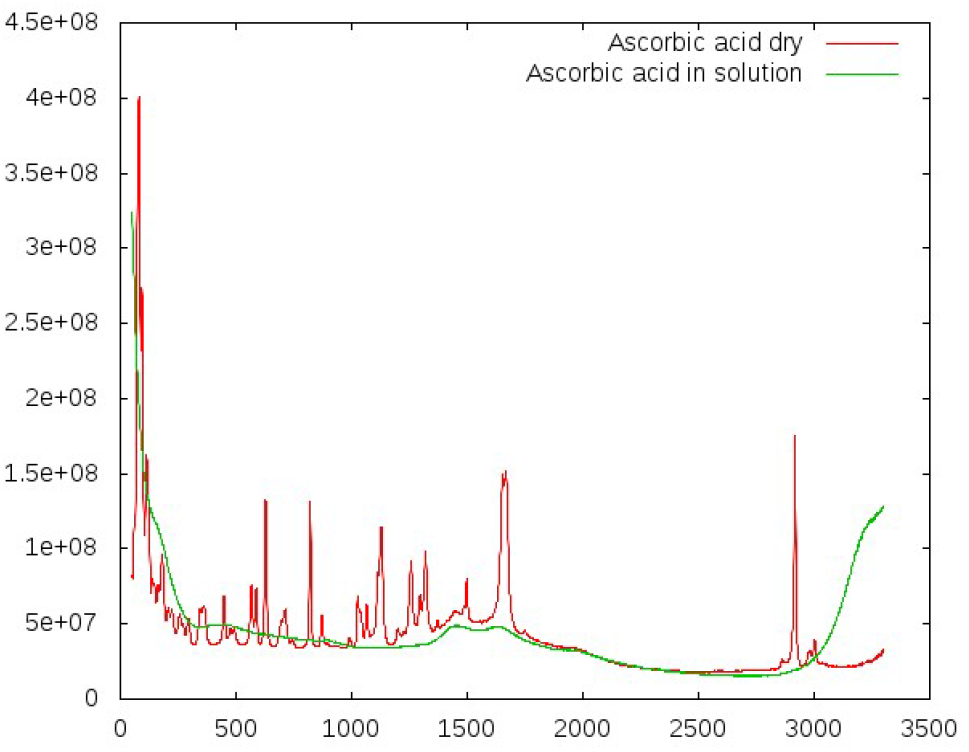
Comparing dry samples and samples in solution: ascorbic acid. All specific peaks vanish in solution for these substances. At around 1500 *cm*^*−*1^, the fluorescence background of the glass carrier is visible. Exposure time: 172 s, concentration: 10 mmol/l.

**Fig 2.**
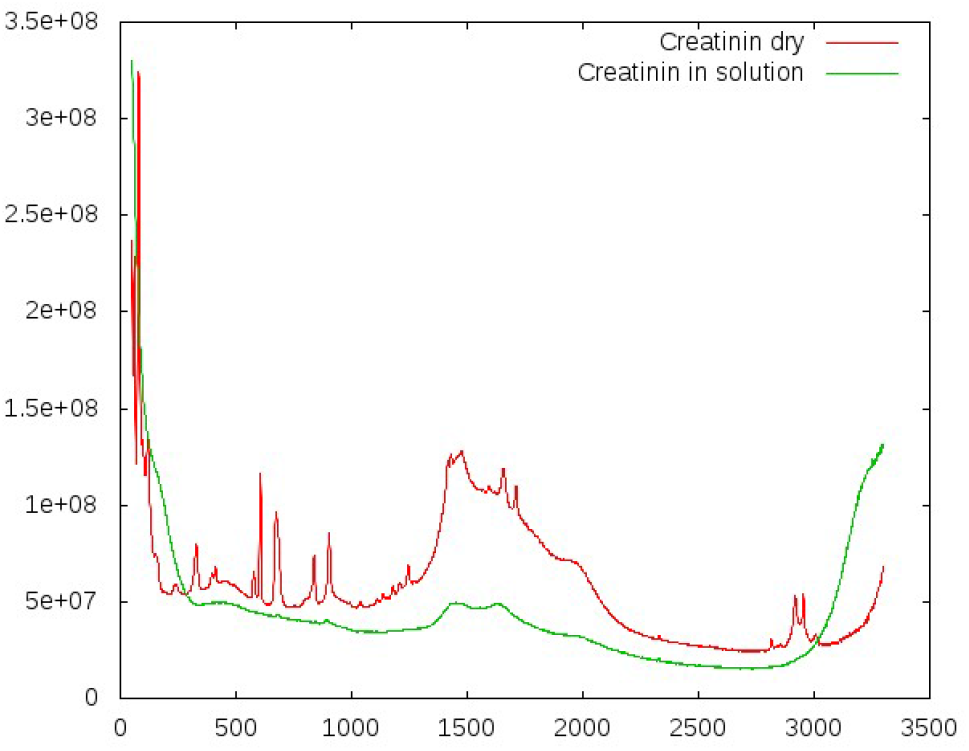
Comparing dry samples and samples in solution: creatinine. All specific peaks vanish in solution for these substances. At around 1500 *cm*^*−*1^, the fluorescence background of the glass carrier is visible. Exposure time: 172 s, concentration: 10 mmol/l.

Information is still contained in the spectra, although it is not as obvious as in the spectra of dry substances. The characteristic Raman peaks are not apparent in the raw spectra, so a crucial step in data analysis is background filtering. Due to the challenging signal-to-noise ratio, many standard methods, such as principle component analysis (PCA), are not immediately applicable.

In the first part of our analysis, we investigate the general information content in the spectra. The main part of the analysis explores the quality of the data in the context of medical diagnostics, namely the correlation of the spectra with the lab results collected for the same patients.

We collected 234 spectra and reached correlations between 0.7 and 0.91 for 19 of the 38 available markers (see Table 2). The correlation values could be easily increased by increasing the number of considered frequencies and a simple weight optimization, but investigations revealed a strong tendency of overfitting. The reported correlations are based on several mechanisms we introduced to counterbalance overfitting.

**Table 2.** Performance analysis for the highly correlating markers. With a minimum weight of 0.2 and 9 spectral vectors. This is background filtered data, with a minimum correlation of 0.6. The significance of the correlation is reported as z-value. The consistency is estimated by the mean value and the standard deviation of the Pearson score for an ensemble of random subsets of 40% size. For each marker, the number of available spectra is reported. Next, the correlation of the values predicted from 238 spectra from an independent cohort with laboratory data is listed, followed by the number of spectra in the validation set. Some of these markers (MELD score, MELDNa score, INR, HBA1C) cannot be detected in serum by standard diagnostics.

| Marker | Pearson | z-value | $P_{40\% \text{mean}/\text{stdev}}$ | $n_{\text{spectra}}$ | Validation | $n_{\text{val}}$ |
| --- | --- | --- | --- | --- | --- | --- |
| Bilirubin | 0.906 | 12.944 | 0.899 / 0.030 | 234 | 0.897 | 328 |
| Conjugated bilirubin | 0.895 | 12.781 | 0.866 / 0.064 | 221 | 0.602 | 52 |
| Total cholesterol | 0.877 | 11.848 | 0.875 / 0.023 | 234 | 0.649 | 206 |
| Unconjugated bilirubin | 0.852 | 11.127 | 0.847 / 0.038 | 221 | 0.770 | 52 |
| MELD score | 0.845 | 10.670 | 0.845 / 0.026 | 231 | 0.810 | 322 |
| Total protein | 0.843 | 10.606 | 0.842 / 0.025 | 234 | 0.642 | 102 |
| LDL cholesterol | 0.839 | 8.084 | 0.837 / 0.037 | 167 | - | 0 |
| MELDNa score | 0.828 | 10.048 | 0.827 / 0.028 | 231 | 0.763 | 275 |
| Unesterified cholesterol | 0.823 | 9.859 | 0.822 / 0.031 | 234 | - | 0 |
| L-lactate dehydrogenase | 0.817 | 6.919 | 0.744 / 0.134 | 162 | 0.265 | 45 |
| Albumin | 0.807 | 9.244 | 0.806 / 0.028 | 234 | 0.624 | 277 |
| INR (International normalized ratio) | 0.806 | 9.241 | 0.803 / 0.043 | 232 | 0.659 | 322 |
| Cholinesterases | 0.796 | 8.836 | 0.796 / 0.027 | 234 | 0.560 | 64 |
| Alpha fetoprotein | 0.756 | 4.886 | 0.545 / 0.277 | 169 | -0.024 | 175 |
| Prealbumin | 0.743 | 6.907 | 0.746 / 0.029 | 234 | - | 0 |
| HDL cholesterol | 0.731 | 3.905 | 0.723 / 0.058 | 167 | - | 0 |
| Calcium | 0.701 | 5.346 | 0.699 / 0.042 | 234 | 0.272 | 272 |
| HBA1C | 0.683 | 2.071 | 0.680 / 0.056 | 165 | 0.250 | 193 |
| Ferritin | 0.657 | 1.061 | 0.653 / 0.072 | 167 | 0.04 | 54 |
| Cystatin c | 0.651 | 3.487 | 0.650 / 0.047 | 234 | - | 0 |
| Iron | 0.647 | 0.124 | 0.646 / 0.058 | 161 | - | 0 |
| Creatinin | 0.637 | 2.950 | 0.635 / 0.064 | 233 | -0.124 | 328 |
| Aspartate transaminase | 0.630 | 2.703 | 0.595 / 0.133 | 234 | -0.057 | 268 |
| Creatinine | 0.623 | 2.448 | 0.625 / 0.056 | 234 | - | 0 |
| Sodium | 0.618 | 2.255 | 0.616 / 0.055 | 233 | 0.166 | 279 |

## Methods

### Experimental setup

#### Raman spectroscopy

We used the Thermo Fisher Scientific DXR SmartRaman Spectrometer (Waltham, Massachusetts, USA) with an excitation wavelength of 780 nm for data collection. Outgoing Raman radiation was detected with a cooled CCD sensor (at 50^*?*^*C*) in the range from 50 to 3500 *cm*^*−*1^. After a period of thorough testing and comparing various settings, we finally chose the settings listed in Table S1 in the supplementary material.

A crucial parameter is the duration and the repetition of the measurements taken by the device. Obviously, a higher number of repetitions would be better than a lower number for statistics and, most importantly, for noise cancellation. However, substantial photo-bleaching was observed, especially in range between 1500 and 3000 *cm*^*−*1^.

The device’s laser can be focused on different heights. We chose a height such that the background created by the carrier was minimal. The focus was slightly above the level of the coverslips.

Throughout the entire analysis, the dominant background was of obvious concern. Figure S1 in the supplementary material shows all 234 unfiltered and unprocessed spectra plotted at once. It would be hard to identify any distinguishing characteristics of the individual curves. Most visible information can be considered background.

#### Blood samples

We selected 234 blood serum samples from well-characterized patients suffering from liver disease. Only residual serum samples from routine diagnostics were retrospectively used. The ethics committee of the University Hospital Leipzig approved the use of residual serum samples and pseudonymized patient data for this study without informed consent (ethical approval 039-14). Accordingly, an additional requirement for informed consent of the patients was waived.

#### Laboratory analysis

Serum parameters were measured on cobas 8000 analysers (Roche, Mannheim, Germany). The international normalized ratio (INR) was determined in citrate plasma using the ACL TOP 700 System (Instrumentation Laboratory, Lexington, USA). Blood cells were measured in EDTA whole blood using the XN 9000 system (Sysmex, Kobe, Japan).

#### Reagents and solutions

The following reagents and solutions were used: fibrinogen 1g (CSL Behring GmbH, Marburg, Germany), mannitol solution 15% (Serumwerk Bernburg, Germany), human serum albumin 20% (low salt, CSL Behring GmbH, Marburg, Germany), Ringer’s solution (Viaflo, Baxter, Deerfield, Illinois, USA), sodium bicarbonate solution 8.4% (B. Braun Melsungen AG, Germany), and L-ascorbic acid (dm-drogerie markt GmbH + Co. KG, Karlsruhe, Germany). Unless specifically stated, dilutions were carried out with deionized water.

All methods were carried out in accordance with relevant guidelines and regulations.

### Evaluation methods

The the spectra were evaluated in two stages. The first consisted of a number of general tests evaluating the application of RS to serum samples. In the second, the correlation of RS with known parameters was analyzed. The former gives a general idea of information content versus noise over the measured range of frequencies. The latter reflects RS’s potential for use as a diagnostic tool. Crucial concepts for both stages of evaluation are the background filtering and correlation analysis.

#### Background filtering

Tests were performed on several datasets: first the raw data, then the background, and last on the background filtered spectra. Background filtering is an essential step to reveal the information content of the spectra. It is clear that the background is dominated by fluorescence, which may itself contain valuable information and will be subject to future investigations. We applied a common asymmetric least square smoothing algorithm [26], [27] that fits a curve to the bottom of fluctuating data.

The algorithm can be tuned to degree to which it will follow local fluctuations. These parameters may be considered the *stiffness* or *flexibility* of the smoothing curves. Examples for two different settings of the regulating parameters are shown in Figure 3 and 4.

**Fig 3.**
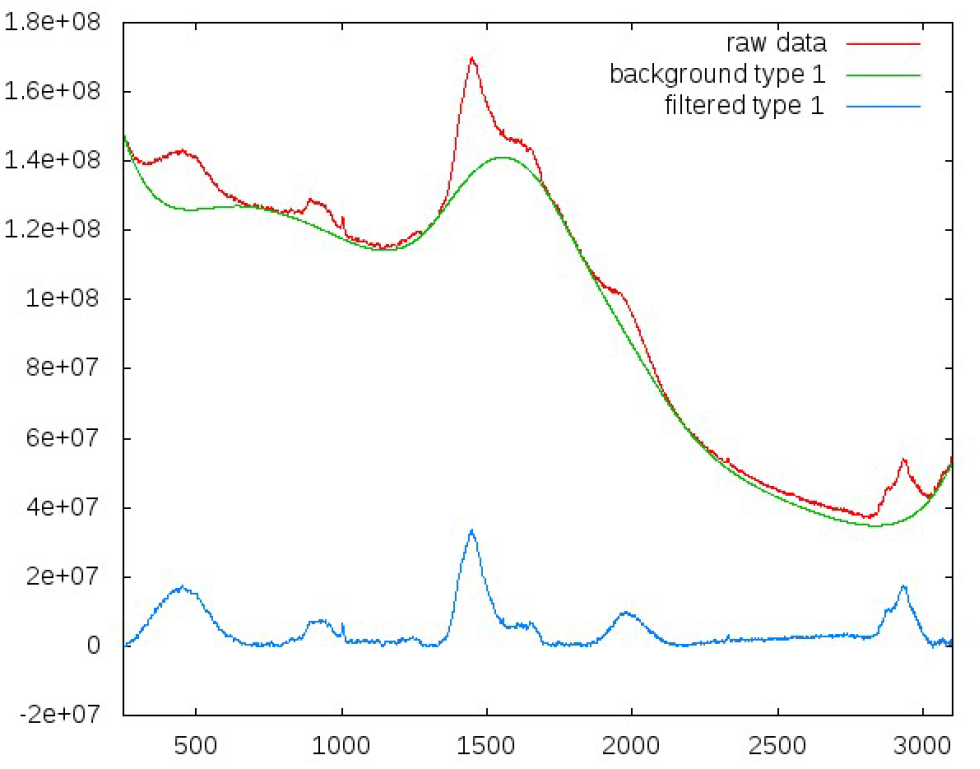
Background filtering with a more ‘rigid’ smoothing curve. Red indicates the original spectra, green the smoothing functions, blue the filtered data.

**Fig 4.**
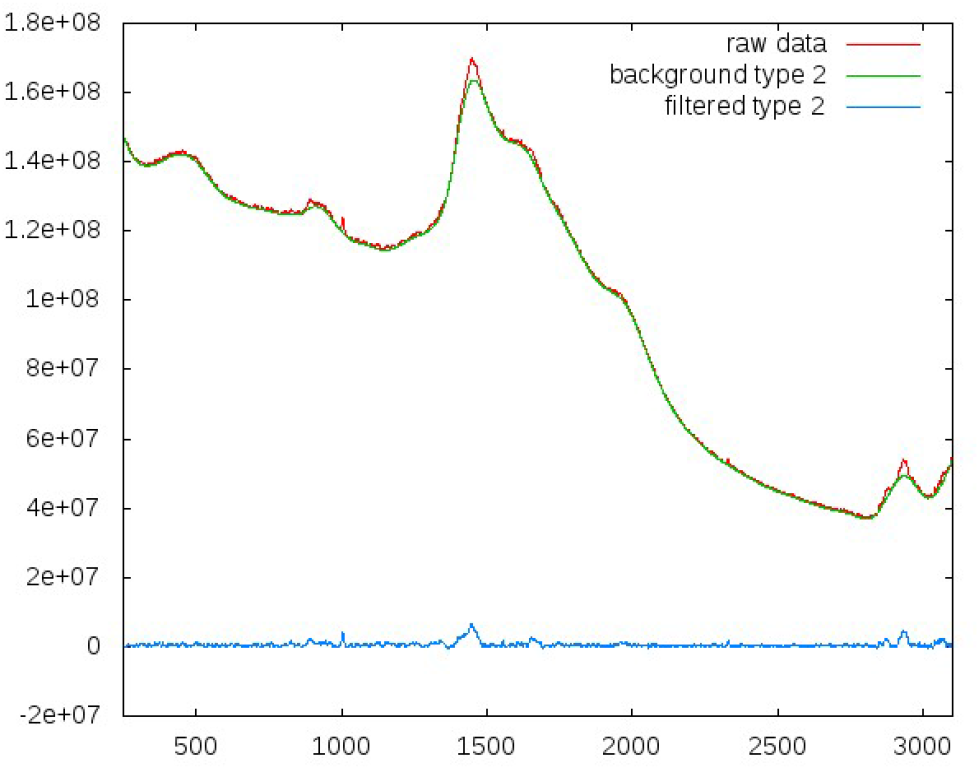
Background filtering with a more ‘flexible’ smoothing curve, leading to stronger filtering. Red indicates the original spectra, green the smoothing functions, blue the filtered data.

#### Correlation and significance

Correlation is the central measure of our analysis. The Pearson-correlation coefficient is a widespread measure for identifying common patterns in data arrays [28].

Correlations are calculated for *spectral vectors*. A spectral vector 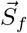 represents a certain frequency *f* and has the size of the number of patients for whom both marker and spectrum are available. The spectra may be considered a *n* ×*k* matrix **S**, where *n* is the number of spectra and *k* the number of frequencies contained in each spectrum. The vector 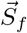 is then the column of matrix **S**, which represents frequency *f* (depicted in red in Figure S1 of the supplementary material.

The starting point of our analysis is the pairwise correlation of a spectral vector 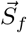 with a marker vector 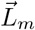, both of dimension *n*. Marker *m* can be any value measured in the medical laboratory for a given patient (e.g. the creatinine level or mortality rate). For a given marker *m* and frequency *f*, the correlation coefficient is given as 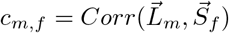.

The final score is the correlation of a marker vector with a linear combination of spectral vectors, which can be written as the product of the matrix **S**, containing all spectra and a weight vector 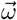. Thus, it can be written as follows:

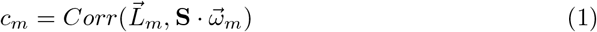

Keeping the number of weights at non-zero low was found to be necessary to avoid overfitting. Additionally, we calculate a z-value as an estimator of the significance of the correlation *c*_*m*_:

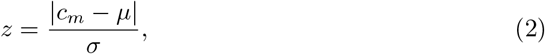

where *µ* is the mean and *σ* the standard deviation of a reference random distribution. The larger z is, the lower the chance of obtaining a given correlation by chance. For every combination of the number of non-zero weights 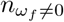 and the number of spectra *n*_*spectra*_, a reference distribution was calculated. For each 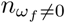 and *n*_*spectra*_, we created 100 random marker vectors 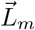. The corresponding correlations were calculated according to the protocol for the markers.

## Results

### General analysis of spectra

As a first test, we investigated the impact of substances being measured in solution rather than being measured dry. We collected spectra both for dry samples and in solution for ascorbic acid and for creatinine (Figure 1 and 2). These revealed a systematic and dramatic issue: all Raman peaks of the solubilized substances vanished completely. Thus, the extraction of relevant information would face a poor signal-to-noise ratio.

Noise can generally be counterbalanced by increasing the sampling, in this case, the number and duration of measurements. However, we observed rather strong frequency-dependent photo bleaching. The effect increased with increased frequency. This is reflected in most spectra; see also Figure S2 of the supplementary material.

As the next qualitative test, we compared the spectra of several pure substances in solution. Figure 5 shows spectra for different single-substance solutions. In some cases, differences in the form of specific peaks are visible to some degree. However, such specific peaks are not visible in the spectra of the serum presented in Figure S1 of the supplementary material. A reason for this could be the very low concentration of proteins and metabolites. The complex molecular mixture of serum does produce some very characteristic vibrational modes, observable in Figure S1 e.g. for the *CH*_2_ − *CH*_3_ stretching modes in the region ranged from 2840 to 3200 *cm*^*−*1^.

**Fig 5.**
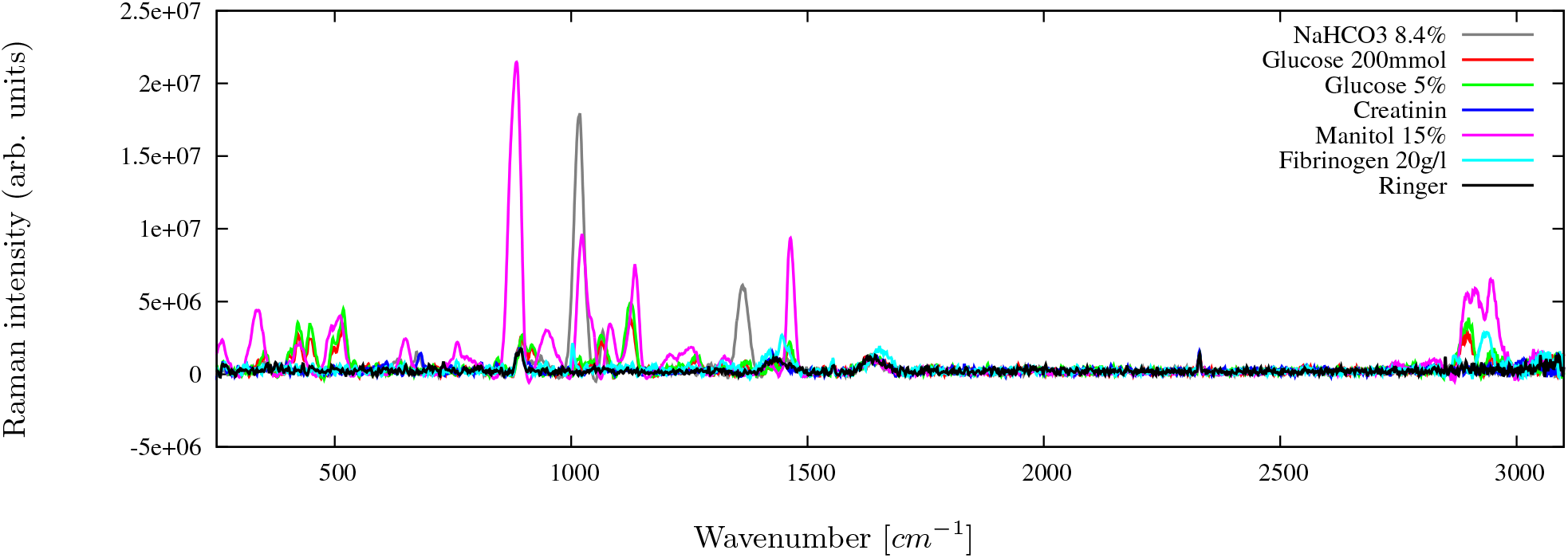
Comparing spectra of individual substances. Shown is the range from 250 to 3100 *cm*^*−*1^. For NaHCO_3_ and mannitol, creatinine specific peaks are visible, as well as for glucose, fibrinogen, although they are less pronounced. This indicates that not all information is lost in solution. Shown are the background filtered spectra. Exposure 172 s.

Albumin is a substance of interest when dealing with liver cirrhosis patients. Figure 6 shows spectra for four different concentrations of albumin, including the absence of albumin. One can distinguish two visible effects: an increasing general offset with increasing concentration caused by fluorescence, and an increasing rise of distinct local peaks due to Raman response. Both effects are already visible in the raw data. After background filtering, these spectra are used as one of the independent tests described below.

**Fig 6.**
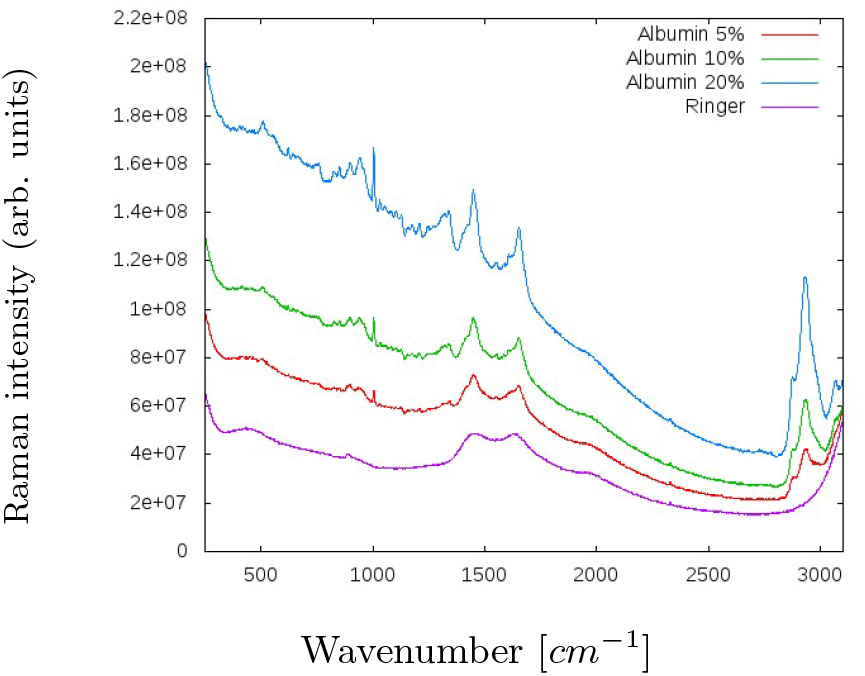
Comparing different albumin concentrations in solution. The spectrum for Ringer solution is plotted as reference. Shown are the unfiltered, raw spectra. The increase of both fluorescence and Raman signal are clearly visible. Exposure 172 s.

The following tests were then performed on the actual serum samples. We collected spectra for each patient six times from the same serum sample and two additional times with 12.5 *µ*l spiked in creatinine and fibrinogen. Figure S2 in the supplementary material shows these for two respective patients. The spectra show a rather large offset in the background. The variation in background between different measurements is greater than differences in Raman intensities. The spiked spectra do not indicate many additive-specific characteristics, such as unique peaks.

### Analysis of spectra versus markers

For every patient for whom we gathered a spectrum, the results of 38 markers were available. The marker values represent concentrations of, for example, albumin or creatinine. Ideally, each of these values could lead to a predictor, meaning that one could predict the creatinine concentration for a patient from a given serum spectrum, for instance. It should be noted that in this study, we investigate RS’s potential to be used as diagnostic tool. The actual optimization of a predictor is not the goal. The distinct correlations between predicted values and laboratory measured values found in this study show clearly that it is possible to derive such predictors. The actual fitting of predictors should, however, be grounded in larger cohorts. One of the main reasons for this is the danger of overfitting, which would level out with larger cohorts.

#### Pairwise correlations

We use the correlation between a marker and a single spectral vector for a given frequency as a first quality estimate. The higher the correlation, the less the frequency reflects the pattern of that marker. In this way, one should be able to identify regions in the spectrum that carry information concerning the marker. Ideally, these frequencies belong to the *hidden* Raman peaks of the substances, revealing themselves by the massive noise. Figure 7 shows the marker with the highest pairwise correlations, bilirubin S. Even in this case, the maximum correlation is only around 0.6, which is far from sufficient. Pairwise correlations alone are an insufficient measure to reflect the markers, which is expected based on the vanishing of peaks when substances were dissolved in solution.

**Fig 7.**
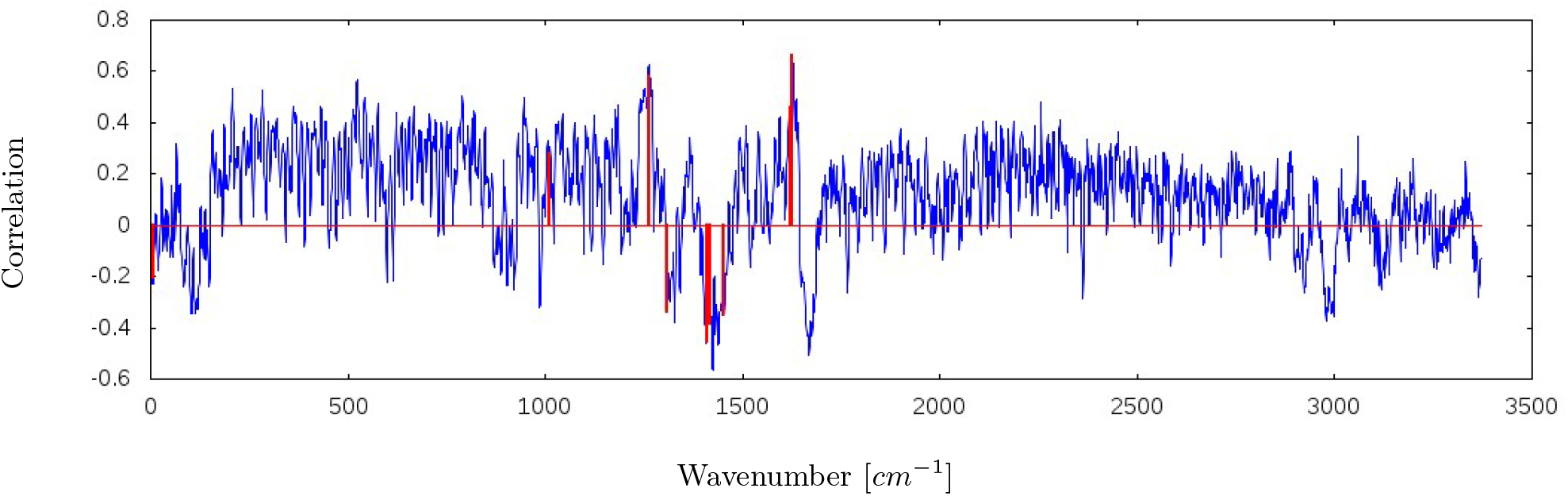
Pairwise correlation between spectral vector 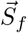 and marker vector 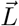 for the marker with the highest correlation, billirubin, is shown in blue. The x-axis shows the columns of the spectral matrix S, representing a frequency *f*. From the pairwise correlations, a linear combination is derived, leading to a improved correlation with the marker while avoiding overfitting. The selection of weights leading to the highest correlation are shown in red. The minimum weight was set to 0.2, and nine spectral vectors were allowed, leading to a final correlation of 0.91 and a z-value of 12,94. The two triplets of nearby spectral vectors may indicate that weight optimization (beyond pairwise values) could lead to a further increase in correlations. Exposure time: 104 s.

#### Overfitting

The most crucial step toward meaningful correlations was to use a linear combination of spectral vectors of different frequencies, according to Formula (1). In our first attempt, we developed an optimizer that switched individual weights from zero to their pairwise correlations, starting from the highest pairwise correlation and accepting any weight that would increase the correlation. Subsequently, we used a Monte Carlo optimizer to fine-tune the weights toward maximum correlation, leading to stellar results: For most markers, we obtained correlations of up to 0.95.

As negative control, we used random vectors instead of markers and performed the same procedure. It was found that even a list of purely random numbers can easily be correlated above 80% using this procedure, rendering the correlations with the markers meaningless.

Using 50 spectral vectors, for example, leads to random correlations of around 85%, with a standard deviation of ∼2%. Single frequencies were not sufficient to represent the markers. The use of too many spectral vectors or too many weights *ω*_*f*_ *≠* 0 led to massive overfitting.

This implied that a significance value was needed (see Equation (2)). At the same time, the number of weights *ω*_*f*_ *≠*0 had to be limited to avoid overfitting. Furthermore, we introduced a minimum value for the pairwise correlations. Frequencies with a pairwise correlation below that minimum would not be considered.

#### Final results

Table 2 lists all markers that reached a final correlation ≥0.6, while restricting the number of spectral vectors to 9. Selecting 9 of 3371 frequencies to construct a basic linear model for up to 234 marker values significantly limits the likelihood of obtaining a result by chance and, to the same extent, avoids overfitting. The table lists the Pearson correlation, the significance z and the number of spectra (markers) that were available for that marker.

As an additional consistency check, we selected 1000x a random subset and then repeated the procedure. For each repetition, around 40% of all pairs of spectra and laboratory values were randomly selected and for this subset the weights were optimized as outlined. Both the mean value and standard deviation of the Pearson correlation obtained by this test are listed in the table as *P*_40%,*mean*_ and *P*_40%,*stdev*_. For a *well-behaving* dataset, the mean should be similar to the Pearson correlation of the full dataset and have a small standard deviation.

Figure 7 shows the pairwise correlation of bilirubin with the spectra, in addition to the selected frequencies, used as spectral vectors, leading to a correlation of 0.91. Weights were not further optimized at this point, so their values reflect the pairwise correlation. The threat of overfitting should have been sufficiently avoided, given the obtained z-values. For comparison, when using nine spectral vectors, bilirubin reaches a z-value of 12.94, and when using 100 spectral vectors, it would reach a z-value of ∼1,38 (assuming this would lead to the same correlation as with nine spectral vectors).

Reducing the number of spectral vectors (*ω*_*f*_ *≠* 0) to just nine is likely to be more strict than necessary. The z-values obtained using 11 spectral vectors do not differ substantially from those obtained using nine. We also performed no weight optimization to avoid overfitting. Larger cohorts would allow for cross-validation, rendering such measures unnecessary. Our goal was to obtain absolutely reliable correlation values. They can be considered the lower threshold of the expected range; the actual correlations will be larger.

### Independent validation

In our first independent test, we collected spectra for different dilutions of an albumin infusion solution (see Figure 6). These spectra were not included when deriving the weights of the spectral vectors.

We used the weights derived from serum spectra to predict the pure albumin concentrations. The results are shown in Table 3. The first column shows the actual content of the sample. Ringer and de-ionized water can be considered to contain 0% albumin. The second column lists the value calculated from spectra using the serum weights. The third column lists the normalized value, which should reflect the albumin concentration. The scaling factor was

**Table 3.** Validating the predictive model with well-defined concentrations of albumin. The predicted value is a linear combination using nine frequencies from the respective spectrum. Both de-ionized water and Ringer solution have an albumin concentration of zero.

| Substance | Predicted value | Scaled [%] |
| --- | --- | --- |
| De-ionized $H_2O$ | -311593.81 | -0.57 |
| Ringer | 52321.53 | 0.096 |
| Albumin 5% | 2896869.41 | 5.34 |
| Albumin 10% | 4851354.12 | 8.94 |
| Albumin 20% | 11251907.89 | 20.73 |

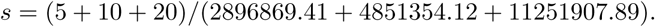

Considering that this is a pilot study with a limited number of patients and that substances in solution show only very weak Raman signals, the agreement between defined and predicted concentrations is already convincing. In addition, the complex compositions of sera are essentially different than pure substances. Thus, the accuracy of the concentration prediction was a convincing confirmation of our approach.

It is noteworthy that the albumin concentrations that were used as validation are well defined concentrations and thus are not as error prone, as it was the case for the laboratory markers used to derive the weights. This agreement supports the correctness of our derivation and, thus, the applicability of RS as diagnostic tool.

For the second test, we analyzed the sera of 11 patients who were not included in the optimization of our method. Figure 8 shows the laboratory values against the calculated values of bilirubin for this data set. A reasonable correlation was observable for most highly correlating markers. However, one spectrum can be considered an outlier for most investigated markers. Interestingly, this spectrum was identified as a *problem case* from the beginning. The serum sample contained a visible residuum, probably from a blood clot. These results further underline the reliability of our strategy.

**Fig 8.**
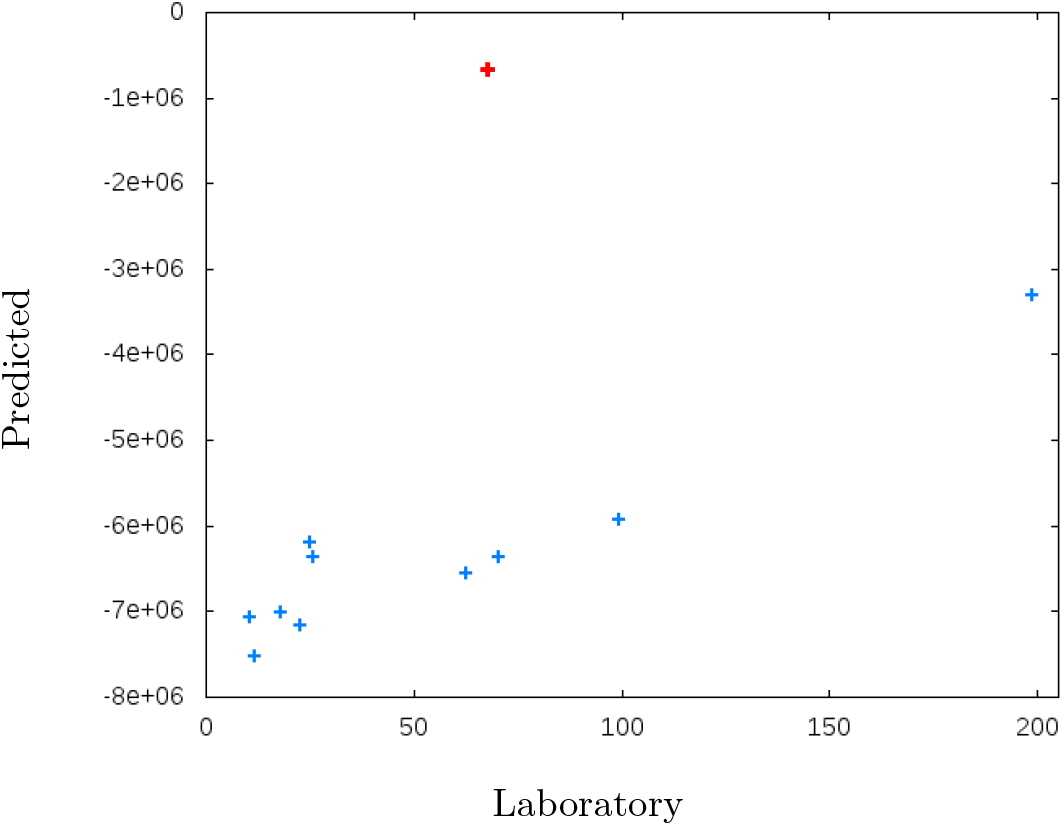
Calculated (y-axis) versus laboratory values (x-axis) for bilirubin of an independent set of 11 patients. The serum sample of the outlier, highlighted in red, contained a visible residuum.

Finally, we completed a large independent validation, using spectra of 328 specimen taken from an evaluation study for liver transplantation in which retention samples were collected for quality assurance. Markers were calculated from the spectra, utilizing the predictive models derived in the main study. Table 2 lists in the ‘Validation’ column the correlations between predicted markers and laboratory values, together with the number of specimen for which laboratory values were available. Several markers reached correlations comparable to the ones reported from the training in the ‘Pearson’ column, which clearly proves the diagnostic value of RS. However, for other markers, the predictions did not reach similar quality. The separation of well validated markers from those that could not be validated may in turn aid analyzing the z-values that were reported in Table 2 for the training data set. The z-values represent a statistical significance estimation for the correlations that should be reflected by the validation in the sense that high z-values should be associated with high correlations in both training and validation. In the case of HBA1C, calcium, L-lactate dehydrogenase, alpha fetoprotein, HDL cholesterol and creatinine the drop in validation correlations is nicely reflected by low z-scores. However, it would be premature at the current state to define a threshold for the z-scores that separates robust correlations from those that are not.

## Discussion

The results of the presented proof of concept study reveal a high potential for RS to be used in medical laboratory diagnostics, although some settings have not yet been systematically optimized. For example, the free parameters in background filtering are currently only a rough approximation, although they are supported by a significant improvement of the correlations.

The development of novel methods to analyze the spectra has great potential for further improvement. However, a detailed theoretical analysis is outside the scope of this paper. The application of standard methods of spectroscopy, such as PCA and spectral deconvolution, would be premature at the current state. The high level of noise renders the *frequency selective linear model* as presented here into being more reliable and thus favorable. Furthermore, it is far from clear that the key assumption of PCA, that a large variation in data corresponds to relevant information, is valid here. Spectral deconvolution remains a longterm goal of this project, which will require both larger cohorts and an exhaustive library of reference spectra of relevant markers in solution at various concentrations.

### Independent tests

The predicted albumin concentrations of the spike experiment are surprisingly accurate, especially when considering that a pure substance is significantly different than the complex molecular mixture that comprises a serum.

In patients with end-stage liver disease, different metabolites and drugs that cannot be detoxified are bound to albumin. The method routinely used in the clinical lab is based on the reaction of albumin with bromocresol green to create a blue green dye, which is measured photometrically [29]. We considered the results of this method to be a gold-standard, aware that relevant influence factors limit the precision of the routine methods, especially in low concentrations [30].

The independent validation study, based on 328 serum samples, clearly reveals the potential of RS to be used as diagnostic tool in the future. It is noteworthy that the spectra were recorded months later and that the specimen come from a different source. The device used for the current study showed significant differences between measurements. The strongest effect was the variation of the background. Another effect that is more profound and more difficult to counterbalance is a frequency shift, due to which Raman resonance peaks will not have their maxima exactly at the expected frequency, but vary slightly from spectrum to spectrum. Although the instrument was calibrated daily as recommended by the manufacturer, frequency shifts of up to 5*cm*^*−*1^ were detectable. This will impede the prediction of markers from spectra to some degree, especially due to our strategy to focus only on a few frequencies in order to avoid overfitting. It is reasonable to assume that improvements of these procedural limitations may allow more robust correlations even for low concentrated markers.

### Results of RS in comparison to clinical diagnostics

Correlations were higher for organic compounds with higher serum concentrations. The most highly concentrated compounds were total protein, albumin, and bilirubin with serum reference concentrations (RC) of 64-83 g/l, 35-52g/l, and *<*10mg/l, respectively. These parameters seem to be valid measurable by means of RS. Bilirubin, which has an intensive color and fluorescence activity, may also improve the correlation, due to still suboptimal background filtering. Prealbumin (RC 0.2-0.4g/l) and AFP (RC *<* 0.007g/l) with lower serum concentrations (0.2-0.4g/l and *<* 0.007g/l, respectively) were less valid measurements.

Identifying markers that are precisely predictable by means of RS would be of obvious benefit. However, it is unlikely that concentration is the only determining factor. Others that might contribute are properties such as size and polarity as well as the molecular environment. This will require extended future research and is out of the scope of this proof-of-concept study.

Compared to liver function markers, the measurements of renal function markers seem to be more sophisticated. Creatinine is the most routinely used biomarker for renal function. Creatinine is a small organic molecule with a molecular mass that has a reference concentration in serum of *<*104*µ*mol/l ∼11.8 mg/l. Due to the small molecular size, only a limited number of specific Raman peaks could be expected, which may be an explanation for the rather low correlation between RS and lab diagnostics. An alternative biomarker is the small protein cystatin C (13.3 kDa, Albumin: ∼66 kDa), which has reference concentrations of 0.47-1.09 mg/l. Due to the low blood concentrations and its small size, its quantification is challenging.

Interestingly, calcium and iron seem to be detectable by RS in physiological concentrations, whereas sodium and chloride revealed only low correlations even though they are more highly concentrated than calcium and iron in serum. Iron and calcium are bound mostly to proteins. This protein interaction may explain the results. The quantification of liver enzymes by RS also achieved limited success. Due to their different detection principles, enzymatic concentrations but not enzymatic activities may be detectable using RS. However, the concentrations of ASAT, ALAT, AP, and gGT are rather small (i.e. ASAT 34*µ*g/l for healthy individuals [31]) and, due to their big molecular weight, molecular concentrations are extremely low.

We also found distinct correlations of RS with glycated hemoglobin (HBA1C) concentration and INR, although these parameters should not be detectable in serum. These findings need some interpretation. The correlations with serum HBA1C results may be explained by the glycation of other serum proteins, such as albumin. The proportion of glycated albumin is known to correlate strongly with that of glycated hemoglobin [32]. Furthermore, in patients with severe disease, the concentration of many molecules will be significantly altered. Therefore, different biomarkers of liver function will be strongly correlated in patients with end-stage liver diseases. Spectra reflect the total composition of serum rather than individual molecules. This may explain the possibility of predicting some of these biomarkers even if they were filtered out beforehand. It has to be emphasized that this of course a pure assumption at this point. However, it might lead to some insights into multi-factorial disease mechanisms. Elucidation of the underlying mechanisms requires an interdisciplinary research approach and is outside the scope of the study objective presented here.

Finally, we were interested in RS’s ability to predict the prognosis of our patients with end-stage liver disease. The most established prognostic laboratory score for these patients is the MELD score. Our analysis showed especially good correlations with the original MELD score without the incorporation of serum sodium concentration, as well as promising results for predicting hepatic encephalopathy and 90-day outcome by RS.

### Outlook

Although the size of our cohort exceeds that of many other studies in this field, the reliability of predictions would immediately benefit from a further increase in cohort size. Given a larger sample size, systematic optimization procedures will most likely improve the signal-to-noise ratio. More precisely, a crucial triangle on which to shed further light is that of exposure time and its relation to photo bleaching versus noise reduction. Different spectrometers will be compared to ensure that repeated measurements under the same conditions lead to identical spectra.

For the subsequent data processing, the background filtering parameters also will require systematic optimization. The application of noise filtering algorithms is likely to be fruitful when used with larger cohorts and optimized parameters. Furthermore, the application of learning algorithms, in combination with our analytic approaches, is expected to boost the prediction quality significantly.

At this state, it remains ambiguous why correlations of some of the analysed markers failed in the validation. It currently cannot be concluded whether these markers are inherently not predictable by means of RS or the presented validation failed due to rectifiable shortcomings. This needs to be clarified in detail in future studies. We are confident that comparing different devices and eliminating technical issues described before will in future also improve the correlations for those markers that failed in this study during the validation.

In addition to optimizing RS data collection and evaluation, we anticipate to explore complementary information from infrared spectroscopy and, potentially, mass spectroscopy.

### Conclusions

In this report, we investigated RS’s potential for application as diagnostic tool for blood serum samples with a cohort of 234 patients at different stages of liver disease. Thirty-eight biomarkers from were available from the clinical laboratory. After establishing a suitable compensation for the background, linear correlation analyses were performed to identify Raman shifts with a reasonable correlation with the clinical laboratory results.

It has to be emphasized that the results should be considered a first estimation. There is a range of technical as well as methodological aspects that require detailed analysis in future studies that have the potential to improve the results. We would expect many more markers to be determinable reliably from spectra. Our study did not attempt to specifically improve biomarker predictions for individual parameters, but rather to develop an algorithm to identify a large number of biomarkers that can potentially be measured by Raman spectroscopy.

It was essential to counterbalance overfitting. Keeping the number of frequencies low was found to be absolutely necessary to avoid overfitting. Relatively few frequencies suffice to obtain good correlations with the markers. From our list of 38 collected parameters, 25 reached a correlation larger than 0.6. The derivation of the weights was performed rather restrictively to strictly avoid overfitting. Thus, the presented correlations, although very good, can be considered conservative approximations. The applied method makes few assumptions about the quality of the data and can be considered more robust than classical approaches, such as PCA. Further studies with larger cohorts will allow for an advanced optimization of the procedure, leading to further increased correlations while preventing overfitting.

It must be emphasized that we did not compare the spectra with error-free reference methods. Rather, we exploited other routine measurements as references. As these measurements contain errors themselves, a correlation above ∼90% would be rather surprising. In clinical lab practice, there are also considerably different results analyzing the same biomarker with different established methods, especially in patients with end-stage liver cirrhosis [33], [34], [35].

We were able to validate our analysis using a defined solution of albumin in different concentrations. Additionally we performed a large scale validation study in which about 10 markers showed robust correlations. However, some, especially lower concentrated markers could not be validated. The results substantiate our methodological approach in general. In summary, the results of our study illustrate the potential of RS to be applied as an untargeted diagnostic tool for many different parameters in clinical laboratory diagnostics.

## Data Availability

All relevant data is included in the manuscript and supplementary material.

## Additional information

The authors declare no competing interests.

## Supplementary material

Table S1: Used settings of the DXR SmartRaman.

Figure S1: Unfiltered spectra of entire cohort of 234 patients. In red is an arbitrary spectral vector 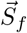 illustrated, for a wavenumber of ∼2000 *cm*^*−*1^.

Figure S2: Reproducibility of spectra from the same sample. Each spectrum shows repeated measurements of a single patient, respectively. The first six curves are independent spectra from the same sample. The last two spectra are taken using the same sample, spiked with 1:1 Creatinine and Fibrinogen solution (final concentrations: creatinine: 5mmol/l, fibrinogen 10g/l). The enrichment was performed to detect specific peaks caused by the additives. However, no specific peak could be assigned from visual inspection. Exposure time: 172s.

