## Supplementary material for "Raman spectroscopy on blood serum samples of patients with end-stage liver disease"

| | |
|---|---|
| Number of sample scans | 30 |
| Collection length | 104.00 sec |
| Number of background scans | 512 |
| Raman laser wavenumber | 12817.90 $cm^{-1}$ |
| Number of data points | 3371 |
| Min/Max/Delta wavenumber | 50.5/3300/0.96 $cm^{-1}$ |

Table S1: Used settings of the DXR SmartRaman.

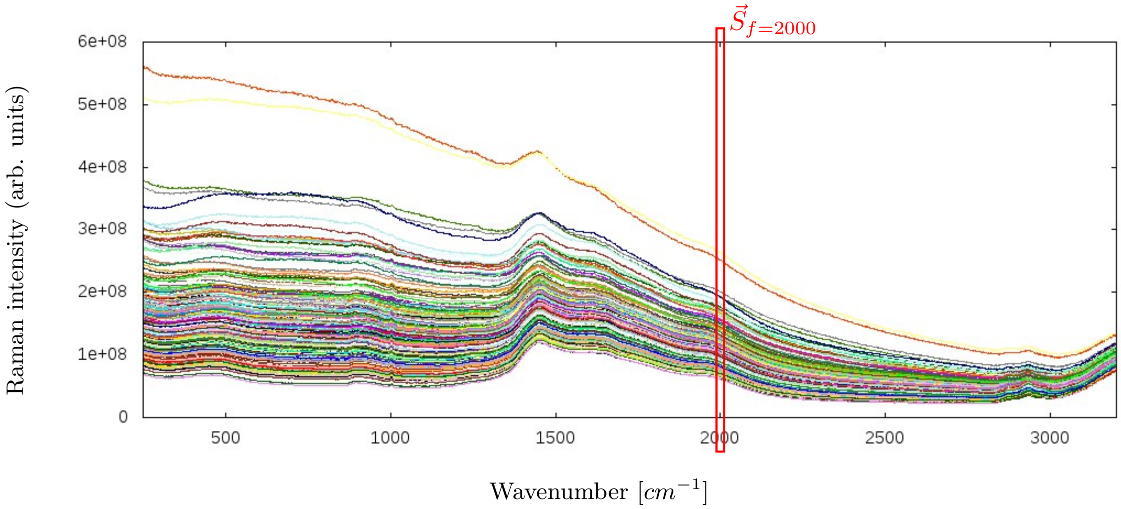


Figure S1: Unfiltered spectra of entire cohort of 234 patients. In red is an arbitrary spectral vector $\vec{S}_i$ illustrated, for a wavenumber of ~2000 $cm^{-1}$.

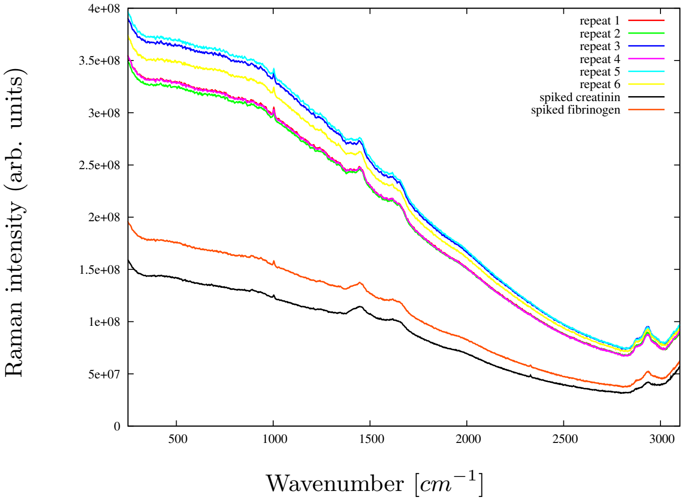


Figure S2: Reproducibility of spectra from the same sample. Each spectrum shows repeated measurements of a single patient, respectively. The first six curves are independent spectra from the same sample. The last two spectra are taken using the same sample, spiked with 1:1 Creatinine and Fibrinogen solution (final concentrations: creatinine: 5mmol/l, fibrinogen 10g/l). The enrichment was performed to detect specific peaks caused by the additives. However, no specific peak could be assigned from visual inspection. Exposure time: 172s.
